## Supplemental tables and figures for "The impact of a demand-side sanitation and hygiene promotion intervention on sustained behavior change and health in Amhara, Ethiopia: a cluster-randomized trial"

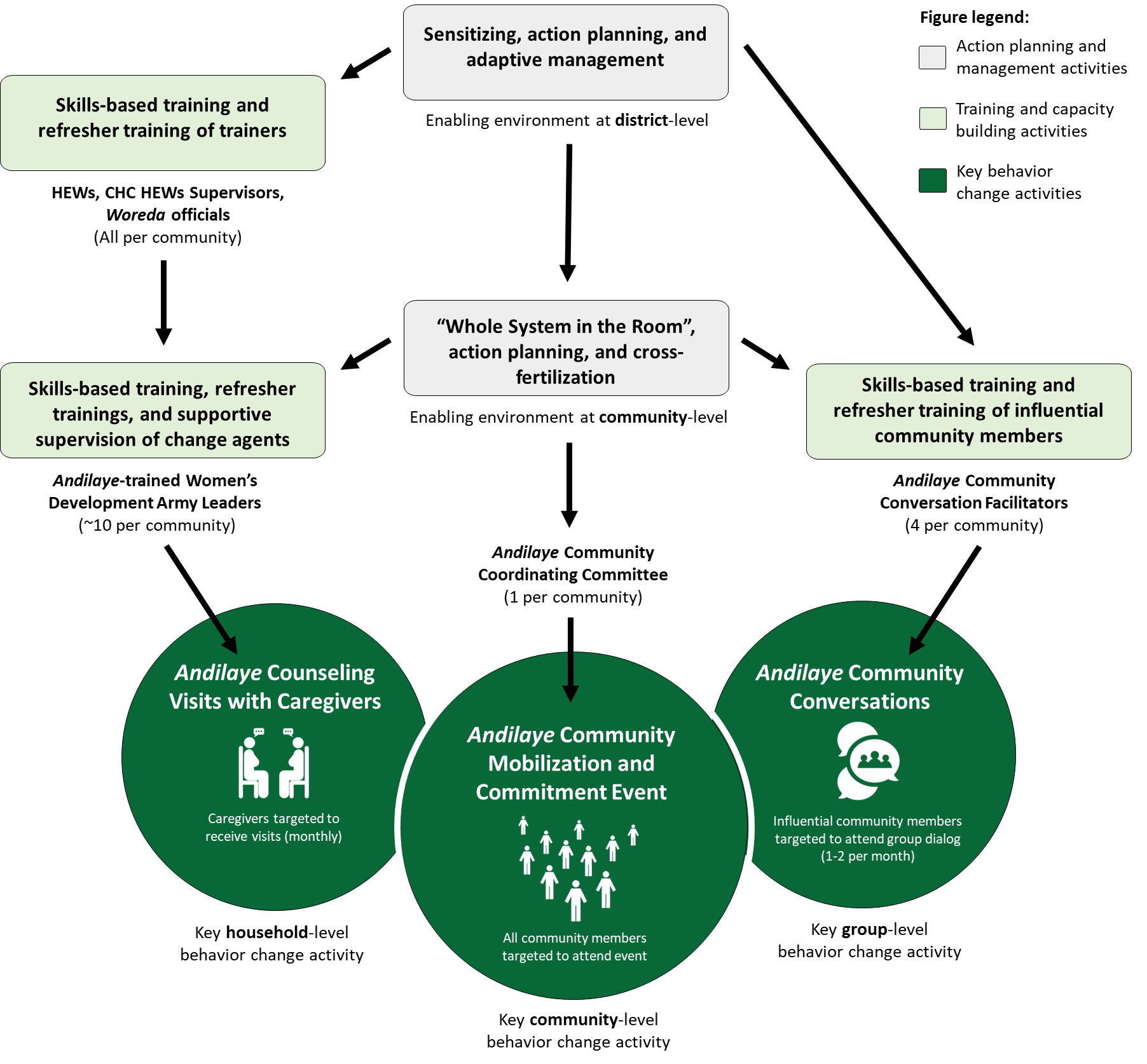

**Supplemental Figure 1.** Diagram summarizing the *Andilaye* intervention

**Supplemental Table 1.** Summary of the *Andilaye* intervention activities and dates of delivery

| **Action planning and management** | **Activity aim** | **Level (type)** | **Dates of delivery ^a^** |
| --- | --- | --- | --- |
| (1) Sensitizing and action planning workshop | To orient key stakeholders to the *Andilaye* intervention and engage them in intervention action planning so as to generate buy-in and foster an enabling environment in which the intervention can be implemented. | District (catalyzing) | September to October 2017 |
| (2) Whole system in the room and action planning | To engage key community stakeholders, orient them to the *Andilaye* intervention, and facilitate their involvement in intervention action planning. This participatory approach aims to generate buy-in and foster an enabling environment (i.e., social opportunity) in which the *Andilaye* intervention can be supported and effectively implemented for a “*strong, caring, healthy community*.” | Community  (catalyzing) | January to  March 2018 |
| (3) Adaptive management workshop | To leverage monitoring data to facilitate evidence-based, controlled, and documented operational-specific modifications during critical program moments (i.e., “change gates”). To improve intervention outcomes and resource management by learning from monitored program outcomes. | District  (maintenance) | February to  March 2019 ^c^ |
| (4) Cross-fertilization visits | To provide an opportunity to share experiences across different intervention communities – to address common implementation bottlenecks, propose solutions, and share perspectives on preliminary behavior change and health outcomes. | Community  (maintenance) | February to  March 2019 ^b^ |
| **Training and capacity building** | **Activity aim** | **Level (type)** | **Dates of delivery ^a^** |
| (1) Skills-based training of the trainers for HEWs, CHC HEWs Supervisors, *Woreda* officials | To provide skills-based training to Health Extension Workers (HEWs), Cluster Health Center (CHC) HEWs Supervisors, and *Woreda* officials on household-level intervention activities, supportive supervision, and on-the-job-training so HEWs can, in turn, effectively train Women’s Development Army Leaders (WDALs) on the implementation of household-level activities and provide supportive supervision. | District  (catalyzing) | December 2017 to January 2018 |
| (2) Skills-based training of WDALs | To provide skills-based training to WDALs on household-level intervention activities, as detailed in the training of the trainers for HEWs, CHC HEWs Supervisors, and *Woreda* officials. | Community  (catalyzing) | January to  February 2018 |
| (3) Training of community conversation facilitators | To provide comprehensive facilitator training to selected *gott* and *kebele* stakeholders on the ‘community conversations’ group-level intervention activity. | District  (catalyzing) | August to  October 2018 ^b^ |
| (4) Skills-based refresher training of the trainers for HEWs, CHC HEWs Supervisors, *Woreda* officials | To reinforce previously acquired knowledge and skills and address trainer/facilitator turnover. Prior experience indicates that such trainings serve to sustain actor motivation and further strengthen capacity. | District  (maintenance) | November 2018 |
| (5) Skills-based review meetings and refresher trainings for WDALs (round 1) | To reinforce previously acquired knowledge and skills, address WDAL turnover, and review successes and address challenges faced in implementing counseling visits with caregivers. Prior experience indicates that such trainings serve to sustain actor motivation and further strengthen capacity. | Community  (maintenance) | November to December 2018 |
| (6) Skills-based review meetings and refresher trainings for WDALs (round 2) | To reinforce previously acquired knowledge and skills, address WDAL turnover, and review successes and address challenges faced in implementing counseling visits with caregivers. Prior experience indicates that such trainings serve to sustain actor motivation and further strengthen capacity. | Community  (maintenance) | February to  March 2019 |
| (7) Skills-based refresher training of community conversation facilitators | To reinforce previously acquired knowledge and skills and address trainer/facilitator turnover. Prior experience indicates that such trainings serve to sustain actor motivation and further strengthen capacity. | District  (maintenance) | February to  April 2019 ^b^ |
| **Key behavior change activities** | **Activity aim** | **Level (type)** | **Dates of delivery ^a^** |
| (1a) Counseling visits with caregivers | To provide personalized counseling to caregivers to equip them with the knowledge, skills, and motivation necessary to adopt improved WASH practices. To foster action capacity, self-efficacy, and barrier planning so caregivers maintain the improved WASH practices. | Household  (catalyzing) | February 2018 to  May 2019 – As dictated by HEWs and WDALs after trainings |
| (1b) Follow-up barrier planning counseling visits with caregivers | To provide continuous follow-up to households such that the house graduates from counseling related to initial adoption of improved practices to counseling related to behavioral maintenance skills. These visits will progressively focus more and more on specific barrier identification and planning skills so the caregiver can maintain his/her improved WASH practices, especially as personal setbacks, systemic shocks, and other obstacles arise. | Household  (maintenance) | As dictated by household progress |
| (2) Community mobilization and commitment event | To improve action knowledge, barrier identification and planning, and attitudes regarding targeted NTD-preventive WASH behaviors through a form of contextually appropriate and interactive edutainment. To initiate the process of shifting social norms through community-generated and managed by-laws and sanctions and public commitment thereof. | Community  (catalyzing) | March to  April 2018 |
| (3a) Community conversations | To change factual beliefs and attitudes, enhance action knowledge, improve perceptions of capability, identify and make plans to overcome barriers, and shift social norms regarding targeted behaviors through community group dialogue**.** To carry out demonstrations that address key factors associated with both breaking away from unimproved practices and adopting improved sanitation and hygiene practices. | Group  (catalyzing) | October 2018 to May 2019 – As dictated by facilitators after trainings |
| (3b) Follow-up community conversations | To generate community-level dialogue regarding nuanced issues associated with maintenance of improved practices and barriers thereof through a follow-up round of community group dialog. To carry-out demonstrations related to behavioral maintenance issues. | Group  (maintenance) | As dictated by group progress |

^a^ Dates reported reflect the overall range in which activities were delivered among all districts and intervention *kebeles*. ^b^ Considerable were delays faced in scheduling with *woreda* officials of Bahir Dar Zuria resulting in a wider range of delivery dates.

**Supplemental Table 2.** Alignment of relevant roles and responsibilities of the Ethiopian Health Extension Programme (HEP) and *Andilaye* Trial

| **Stakeholder** | **Relevant HEP roles and responsibilities ^a^** | **Alignment with Andilaye Trial** | |
| --- | --- | --- | --- |
| **The Federal Ministry of Health** | **Design:** Determine overall program concept, standards, and implementation guidelines and provide communication tools and materials | **✔** | Engaged during formative research and intervention design |
|  | **Implementation (National-level):** Mobilize national and international resources | **✘** | National implementation and reporting roles and responsibilities were not included as rolling out a government-sponsored program in a select number of areas was both politically and logistically infeasible |
|  | **Reporting:** Establish the health management information system |  |  |
| **The Regional Health Bureaus/Zonal Health Departments** | **Design:** Adapt implementation guidelines to local conditions and communication tools; and adapt materials into local languages and distributes them to woredas | **✔** | Engaged during formative research and intervention design |
|  | **Implementation (Regional/Zonal-level):** Provide technical and administrative support to Woreda Health Offices | **✘** | Regional/Zonal implementation and reporting roles and responsibilities were performed by the Ethiopian-based study team |
|  | **Reporting:** Obtain reports from Woreda Health Offices and provide information to the Ministry of Health |  |  |
| **The Woreda Health Office** | **Design:** Adapt communication tools and materials | **✔** | Engaged during formative research and intervention design |
|  | **Implementation (District-level):** Provide technical, administrative, and financial support to health centers and health posts; and plan and provide in-service training to HEWs and Woreda Health Office staff | **✔** | While financial and overall technical support was provided by the Andilaye team, the Woreda Health Office was designed as the key stakeholder in implementing all district-level activities |
|  | **Supportive supervision:** Provide supportive supervision of HEWs and the overall management of health centers and health posts | **✔** | Supportive supervision and on-the-job-training of HEWs were designed to be completed by HEWs Supervisors via an Andliaye-specific checklist |
|  | **Reporting:** Obtain reports from health posts and health centers and provide information to Regional Health Bureaus or Zonal Health Departments | **✔** | While detailed process data was collected by the Ethiopian-based study team, program monitoring mechanisms were included in the design |
| **Health Extension Workers (HEWs)** | **Implementation (Community-level):**  Manage operations of health posts; and identify, train, and collaborate with WDALs teams | **✔** | While overall technical support was provided by the Ethiopian-based study team, the HEW was designed as the key stakeholder in implementing all community-level activities and engaging additional community change agents for group-level and household-level activities |
|  | **Implementation (Group-level):** Communicate health messages by involving the community social networks, associations, religious institutions, and government structures | **✔** | Over-extension of HEWs was addressed through the engagement of additional community change agents for group-level activities (i.e., community conversation facilitators) and household-level activities (i.e., WDALs) |
|  | **Implementation (Household-level):** Conduct home visits and outreach services to promote preventive actions; and prioritize households with low performance in implementing the package and support them in all the Health Extension Packages that are relevant to them |  |  |
|  | **Supportive supervision:** Provide supportive supervision and evaluation of WDAL teams; and conduct biweekly meetings to evaluate the performance of the development teams | **✔** | Supportive supervision and on-the-job-training of WDALs was designed to be completed by HEWs via an Andliaye-specific checklist |
|  | **Reporting:** Provide reports to Woreda Health Offices | **✔** | While detailed process data was collected by the Ethiopian-based study team, program monitoring mechanisms were included in the design |
| **Women’s Development Army Leaders (WDALs)** | **Implementation (Community, group, and household-level):** Volunteers not institutionalized into the health system; they are regarded as community representatives and reflect government efforts to devolve responsibilities for  health to individuals and local communities by mobilizing the population and supporting HEWs | **✔** | In addition to supporting community and group-level activities, WDALs were trained to provide inter-personal counseling with caregivers by conducting Andilaye household counseling visits with each household in her catchment area |

^a^ Workie, N.W. and R., Gandham NV, 2013. Ethiopia - The health extension program in Ethiopia, Universal Health Coverage (UNICO) studies series. World Bank, Washington, DC.

**Supplemental Table 3.** Process data for *Andilaye* intervention activities

**Action planning and management activities**

(1) Sensitizing and action planning workshop (district-level catalyzing activity)

| **Process data** | | **Bahir Dar Zuria** | **Fogera** | **Farta** | **Overall** |
| --- | --- | --- | --- | --- | --- |
| **Dose delivered** | Proportion of districts with activity implemented (Sept-Oct 2017) | 1/1 (100%) | 1/1 (100%) | 1/1 (100%) | 3/3 (100%) |
|  | Proportion of districts with activity objectives completed as planned (according to activity checklist) | 1/1 (100%) | 1/1 (100%) | 1/1 (100%) | 3/3 (100%) |
|  | Length of activity (one day workshop) | 8 hours | 7.5 hours | 4.75 hours ^a^ | 6.75 hours |
| **Participation** | Total number of participants | 19 ^b^ | 12 | 16 | 47 |
|  | Proportion of districts with Amhara Regional Health Bureau OR COWASH representative in attendance during workshop | 1/1 (100%) | 1/1(100%) | 0/1 (0%) | 2/3 (66%) |
|  | Proportion of districts with Woreda Health Office representative AND Hygiene and Sanitation Officer in attendance during workshop | 1/1 (100%) | 1/1 (100%) | 1/1 (100%) | 3/3 (100%) |
|  | Proportion of intervention kebeles with administrator in attendance | 7/8 (88%) ^c^ | 6/6 (100%) | 11/11 (100%) | 24/25 (96%) |
| **Dose received** | Proportion of districts with endorsed action plan for the completion of district-level activities and the ‘ Whole system in the room and action planning’ community-level activity and accompanying supportive supervision | 1/1 (100%) | 1/1 (100%) | 1/1 (100%) | 3/3 (100%) |
|  | Proportion of participants that endorsed action plan for the completion of district-level activities and the ‘ Whole system in the room and action planning’ community-level activity and accompanying supportive supervision | 15/19 (79%) | 12/12 (100%) | 15/16 (94%) | 42/47 (89%) ^d^ |
| **Context** | ^a^ Participants had another woreda-level governmental workers’ meeting with the woreda administrator and asked to finish this workshop early. The action plan was supposed to be first completed in two groups and then each group presenting to all participants to finalize. To accommodate the other meeting, the participants completed the action plan as one group and finalized.  ^b^ The workshop completed in Bahir Dar City (the regional capital city) was the one with the highest stakeholder turnout, including regional level government and non-government organizations stakeholders.  ^c^ One Bahir Dar Zuria kebele administrator from a non-study kebele (Wonjeta) was invited by mistake instead the kebele administrator from an intervention kebele (Wondata). Similarity of names created the mismatch while extending invitations.  ^d^ Some workshop participants (mainly regional representatives and woreda administrators) from each of the three districts did not attend the afternoon session of the workshop; this might have impacted the number of action plan endorsing stakeholders. | | | | |

(2) Whole system in the room and action planning (community-level catalyzing activity)

| **Process data** | | **Bahir Dar Zuria** | **Fogera** | **Farta** | **Overall** |
| --- | --- | --- | --- | --- | --- |
| **Dose delivered** | Proportion of intervention kebeles with activity implemented (Jan-Mar 2018) | 8/8 (100%) | 6/6 (100%) | 11/11 (100%) | 25/25 (100%) |
|  | Proportion of intervention kebeles with activity objectives completed as planned (according to activity checklist) | 8/8 (100%) | 6/6 (100%) | 11/11 (100%) | 25/25 (100%) ^a^ |
|  | Average length of activity (partial day workshop) | 1.75 hours | 2 hours | 1.33 hours | 1.75 hours |
| **Participation** | Total number of participants | 64 | 42 | 117 | 223 |
|  | Proportion of intervention kebeles with administrator acting as activity facilitator | 7/8 (88%) | 6/6 (100%) | 11/11 (100%) | 24/25 (96%) |
|  | Proportion of intervention kebeles with Woreda Health Office representatives acting as activity co-facilitator | 1/8 (13%) | 0/6 (0%) | 1/11 (9%) | 2/25 (8%) |
|  | Proportion of intervention kebeles with manager in attendance | 7/8 (88%) | 6/6 (100%) | 9/11 (82%) | 22/25 (88%) |
|  | Proportion of intervention kebeles with at least one Health Extension Worker (HEW) in attendance | 8/8 (100%) | 6/6 (100%) | 11/11 (100%) | 25/25 (100%) |
|  | Proportion of intervention kebeles with at least one Agriculture Extension Worker (AEW) in attendance | 7/8 (88%) | 6/6 (100%) | 10/11 (91%) | 23/25 (92%) |
|  | Proportion of intervention kebeles with at least one school director in attendance | 5/8 (63%) | 5/6 (83%) | 5/11 (45%) | 15/25 (60%) |
|  | Proportion of intervention kebeles with at least one Women’s Development Army Leader (WDAL) in attendance | 4/8 (50%) | 1/6 (17%) | 7/11 (64%) | 12/25 (48%) |
|  | Proportion of intervention kebeles with at least one religious leader in attendance | 7/8 (88%) | 4/6 (67%) | 10/11 (91%) | 21/25 (84%) |
|  | Proportion of intervention kebeles with at least one influential elders or other influential people from the gott | 8/8 (100%) | 4/6 (67%) | 11/11 (100%) | 22/25 (88%) |
| **Dose received** | Proportion of intervention kebeles with Andilaye community conversation facilitators identified in the action plan | 8/8 (100%) | 6/6 (100%) | 11/11 (100%) | 25/25 (100%) |
|  | Proportion of intervention kebeles with a coordinating committee for the ‘Community mobilization and commitment event’ identified in the action plan | 8/8 (100%) | 6/6 (100%) | 11/11 (100%) | 25/25 (100%) |
|  | Proportion of intervention kebeles with a master of ceremony for the ‘Community mobilization and commitment event’ identified in the action plan | 8/8 (100%) | 6/6 (100%) | 11/11 (100%) | 25/25 (100%) |
| **Context** | ^a^ The objectives were completed as planned, however, the expectation of payment (per diem) associated with the activity was an issue and in some cases a clear disappointment for participants. Although this was a one to two hour activity and government policies were followed, participants (a composition of community members and salaried government workers) seem to expect some kind of per diem and when they found out that there is no per diem (at the end of the activity), some participants expressed their disappointment. | | | | |

(3) Adaptive management workshop and cross-fertilization visits (district and community-level maintenance activities)

| **Process data** | | **Bahir Dar Zuria** | **Fogera** | **Farta** | **Overall** |
| --- | --- | --- | --- | --- | --- |
| **Dose delivered** | Proportion of districts with activity implemented (Feb-Mar 2019) | 1/1 (100%) | 1/1 (100%) | 1/1 (100%) | 3/3 (100%) ^a^ |
|  | Proportion of districts with activity objectives completed as planned (according to activity checklist) | 1/1 (100%) | 1/1 (100%) | 1/1 (100%) | 3/3 (100%) |
|  | Length of activity each day (two day workshop) | 8 hours | 8 hours | 8 hours | 8 hours |
| **Participation** | Total number of participants | 25 | 18 | 28 | 71 |
|  | Proportion of districts with Woreda Health Office Head, Hygiene and Sanitation Officer, AND HEW Program Officer in attendance | 1/1 (100%) | 1/1 (100%) | 1/1 (100%) | 3/3 (100%) |
|  | Proportion of intervention kebele Cluster Health Centers (CHC) Heads AND CHC HEWs Supervisors in attendance | 7/7 (100%) | 4/4 (100%) | 7/7 (100%) | 18/18 (100%) |
|  | Proportion of intervention kebeles with administrator in attendance | 8/8 (100%) | 6/6 (100%) | 11/11 (100%) | 25/25 (100%) |
| **Dose received** | Proportion of districts that identified implementation challenges and developed action plan for ways forward for the completion of district, community, group, and household-level activities | 1/1 (100%) | 1/1 (100%) | 1/1 (100%) | 3/3 (100%) ^b^ |
| **Context** | ^a^ Cross fertilization visits were conducted on the second day where all participants visited ‘better performing’ intervention kebeles (as selected on during the first day of the adaptive management workshop). During the visit, participants observed WDALs performing household visits and facilitators conducting community conversations. After observing these activities, the participants came together to further discuss ways forward to overcome implementation challenges in their respective communities.  ^b^ The overall lack of ownership of project activities and existing gaps in implementing the Health Extension Program (HEP) (e.g., supportive supervision and WDAL structure) were major implementation challenge discussed in all districts. | | | | |

**Training and capacity building activities**

(1) Skills-based training of the trainers for HEWs, CHC HEWs Supervisors, *Woreda* officials (district-level catalyzing activity)

| **Process data** | | **Bahir Dar Zuria** | **Fogera** | **Farta** | **Overall** |
| --- | --- | --- | --- | --- | --- |
| **Dose delivered** | Proportion of districts with activity implemented (Dec 2017-Jan 2018) | 1/1 (100%) | 1/1 (100%) | 1/1 (100%) | 3/3 (100%) |
|  | Proportion of districts with activity objectives completed as planned (according to activity checklist) | 1/1 (100%) | 1/1 (100%) | 1/1 (100%) | 3/3 (100%) |
|  | Length of activity each day (two day training) | 10 hours | 10 hours | 10 hours | 10 hours |
| **Participation** | Total number of participants | 38 | 27 | 44 ^a^ | 108 |
|  | Proportion of districts with Woreda Health Office representative AND Hygiene and Sanitation Officer in attendance during training | 1/1 (100%) | 1/1 (100%) | 1/1 (100%) | 3/3 (100%) ^b^ |
|  | Proportion of intervention kebele CHC HEWs Supervisors trained | 7/7 (100%) | 4/4 (100%) | 7/7 (100%) | 18/18 (100%) |
|  | Proportion of intervention kebele HEWs trained | 21/21 (100%) | 15/15 (100%) | 20/21 (95%) | 56/57 (98%) ^b^ |
|  | Proportion of intervention kebeles with at least one HEW trained | 8/8 (100%) | 6/6 (100%) | 11/11 (100%) | 25/25 (100%) |
| **Dose received** | Proportion of intervention kebeles with endorsed action plan for the completion of household-level activities and accompanying supportive supervision | 8/8 (100%) | 6/6 (100%) | 11/11 (100%) | 25/25 (100%) |
| **Context** | ^a^ Two training sessions were conducted in Farta given the high number of intervention kebeles and target participants.  ^b^ A “mop up” training was conducted in Bahir Dar City after the district-level trainings to address 10 HEWs (6 from Bahir Dar Zuria, 2 from Fogera, and 2 from Farta) and Bahir Dar Zuria Woreda Health Office representative and Hygiene and Sanitation Officer that were absent during the original trainings. | | | | |

(2) Skills-based training of WDALs (community-level catalyzing activity)

| Process data | | Bahir Dar Zuria | Fogera | Farta | Overall |
| --- | --- | --- | --- | --- | --- |
| Dose delivered | Proportion of intervention *kebeles* with activity implemented (Jan-Feb 2018) | 8/8 (100%) | 6/6 (100%) | 11/11 (100%) | 25/25 (100%) |
|  | Proportion of intervention *kebeles* with activity objectives completed as planned (according to activity checklist) | 8/8 (100%) | 6/6 (100%) | 11/11 (100%) | 25/25 (100%) |
|  | Length of activity each day (two day training) | 4.75 hours | 4.0 hours | 7 hours | 6 hours |
| Participation | Proportion of intervention *kebeles* with HEWs acting as activity training facilitator | 8/8 (100%) | 6/6 (100%) | 11/11 (100%) | 25/25 (100%) |
|  | Proportion of intervention *kebeles* with CHC HEWs Supervisors acting as activity training co-facilitator | 6/8 (75%) | 6/6 (100%) | 8/11 (73%) | 20/25 (80%) ^a^ |
|  | Proportion of WDALs from intervention *kebeles* in attendance during the training (8-10 per *kebele*) | 72/73 (99%) | 54/56 (96%) | 107/110 (97%) | 233/239 (97%) |
|  | Proportion of intervention *kebeles* with *Andilaye* trained WDALs | 8/8 (100%) | 6/6 (100%) | 11/11 (100%) | 25/25 (100%) |
| Dose received | Proportion of intervention *kebeles* with HEWs reporting that they have received *Andilaye* supportive supervision from CHC HEWs Supervisors after training [during quarterly monitoring surveys in July 2018] | 0/8 (0%) | 2/6 (33%) | 0/11 (0%) | 2/25 (8%) ^b^ |
|  | Proportion of randomly sampled WDALs reporting that they have received *Andilaye* supportive supervision from HEWs after training [during quarterly monitoring surveys in July 2018] | 12/29 (41%) | 4/23 (17%) | 21/42 (50%) | 37/94 (39%) ^b^ |
| Context | ^a^ Training of WDALs was planned to primarily be facilitated by HEWs with help from CHC HEWs Supervisors, but the involvement of CHC HEWs Supervisors was minimal in some *kebeles*. Thus, the Ethiopian-based study team assisted trainers and trained WDALs together with HEWs*.*  ^b^ It was recommended that CHC HEWs Supervisors perform supportive supervision on a routine basis (at least once per month for each HEW), in accordance with the woreda's *Andilaye* action planning document. Similarly, it was recommended that HEWs perform supportive supervision with each WDAL in her catchment area every month, in accordance with the HEW's *Andilaye* action planning document. When conducting supervisory visits, HEWs were trained to use the supportive supervision and on-the-job checklist. However, quarterly monitoring surveys conducted 5-6 months after the training of WDALs (July 2018) indicated that few HEWs have received supportive supervision and on-the-job training from CHC HEW Supervisors and that few WDALs have received supportive supervision and on-the-job training from HEWs. The review of the importance of providing supportive supervision to both HEWs and WDALs was prioritized during ‘Skills-based refresher training of the trainers for HEWs, CHC HEWs Supervisors, Woreda officials’ (Nov 2018) and during ‘Skills-based review meetings and refresher trainings for WDALs (round 1)’ (Nov-Dec 2018). | | | | |

(3) Training of community conversation facilitators (district-level catalyzing activity)

| **Process data** | | **Bahir Dar Zuria** | **Fogera** | **Farta** | **Overall** |
| --- | --- | --- | --- | --- | --- |
| **Dose delivered** | Proportion of districts with activity implemented (Aug-Oct 2018) | 1/1 (100%) | 1/1 (100%) | 1/1 (100%) | 3/3 (100%) |
|  | Proportion of districts with activity objectives completed as planned (according to activity checklist) | 1/1 (100%) | 1/1 (100%) | 1/1 (100%) | 3/3 (100%) |
|  | Length of activity each day (two day training) | 8.33 hours | 8.75 hours | 7.75 hours | 8.25 hours |
| **Participation** | Total number of participants | 34 ^a^ | 26 | 45 ^a^ | 105 |
|  | Proportion of districts with *Woreda* Health Office representative AND Hygiene and Sanitation Officer in attendance | 1/1 (100%) | 0/1 (0%) | 1/1 (100%) | 2/3 (66%) |
|  | Proportion of *Andilaye* community conversation facilitators in attendance | 32/32 (100%) | 24/24 (100%) | 43/44 (98%) | 99/100 (99%) |
|  | Proportion of intervention *kebeles* with at least at least one facilitator per pair of facilitators trained on *Andilaye* community conversations | 8/8 (100%) | 6/6 (100%) | 11/11 (100%) | 25/25 (100%) |
| **Dose received** | Proportion of intervention *kebeles* with representatives that endorsed action plan for the completion of group-level activities | 8/8 (100%) | 6/6 (100%) | 11/11 (100%) | 25/25 (100%) ^b^ |
| **Context** | ^a^ Two training sessions were conducted in Bahir Dar Zuria and Farta given the high number of intervention *kebeles* and target participants.  ^b^ During action planning, it was nearly unanimous that refreshments would need to be provided in order to motivate community members to attend community conversations, as planned. While this was not originally budgeted, each intervention *kebele* was provided coffee and flour for bread to accommodate a coffee ceremony during the community conversations. | | | | |

(4) Skills-based refresher training of the trainers for HEWs, CHC HEWs Supervisors, *Woreda* officials (district-level maintenance activity)

| **Process data** | | **Bahir Dar Zuria** | **Fogera** | **Farta** | **Overall** |
| --- | --- | --- | --- | --- | --- |
| **Dose delivered** | Proportion of districts with activity implemented (Nov 2018) | 1/1 (100%) | 1/1 (100%) | 1/1 (100%) | 3/3 (100%) |
|  | Proportion of districts with activity objectives completed as planned (according to activity checklist) | 1/1 (100%) | 1/1 (100%) | 1/1 (100%) | 3/3 (100%) |
|  | Length of activity each day (two day training) | 8 hours | 8 hours | 8 hours | 8 hours |
| **Participation** | Total number of participants | 29 | 26 | 46 ^a^ | 101 |
|  | Proportion of study districts with *Woreda* Health Office representative AND Hygiene and Sanitation Officer in attendance during training | 0/1 (0%) | 1/1 (100%) | 1/1 (100%) | 2/3 (66%) |
|  | Proportion of intervention *kebele* CHC HEWs Supervisors trained | 7/7 (100%) | 4/4 (100%) | 5/7 (88%) | 16/18 (89%) |
|  | Proportion of intervention *kebele* HEWs trained | 16/21 (100%) | 13/15 (88%) | 25/25 (100%) | 54/61 (89%) |
|  | Proportion of intervention *kebeles* with at least one HEW trained | 8/8 (100%) | 6/6 (100%) | 11/11 (100%) | 25/25 (100%) |
| **Dose received** | Proportion of intervention *kebeles* that identified implementation challenges and developed action plan for ways forward for the completion of household-level activities and accompanying supportive supervision | 8/8 (100%) | 6/6 (100%) | 11/11 (100%) | 25/25 (100%) |
| **Context** | ^a^ Two training sessions were conducted in Farta given the high number of intervention *kebeles* and target participants. | | | | |

(5) Skills-based review meeting and refresher training for WDALs (round 1) (community-level maintenance activity)

| **Process data** | | **Bahir Dar Zuria** | **Fogera** | **Farta** | **Overall** |
| --- | --- | --- | --- | --- | --- |
| **Dose delivered** | Proportion of intervention *kebeles* with activity implemented (Nov-Dec 2018) | 8/8 (100%) | 6/6 (100%) | 11/11 (100%) | 25/25 (100%) |
|  | Proportion of intervention *kebeles* with activity objectives completed as planned (according to activity checklist) | 8/8 (100%) | 6/6 (100%) | 11/11 (100%) | 25/25 (100%) |
|  | Length of activity (partial day training) | 2.75 hours | 3.0 hours | 3.25 hours | 3 hours |
| **Participation** | Proportion of intervention *kebeles* with HEWs acting as activity training facilitator | 8/8 (100%) | 6/6 (100%) | 11/11 (100%) | 25/25 (100%) |
|  | Proportion of intervention *kebeles* with CHC HEWs Supervisors acting as activity training co-facilitator | 7/8 (88%) | 6/6 (100%) | 10/11 (91%) | 23/25 (92%) ^a^ |
|  | Proportion of WDALs from intervention *kebeles* in attendance during the training (8-10 per *kebele*) | 70/74 (95%) | 62/62 (100%) | 104/107 (97%) | 236/243 (97%) ^b^ |
|  | Proportion of intervention *kebeles* with *Andilaye* trained WDALs | 8/8 (100%) | 6/6 (100%) | 11/11 (100%) | 25/25 (100%) |
| **Dose received** | Proportion of intervention *kebeles* with HEWs reporting that they have received *Andilaye* supportive supervision from CHC HEWs Supervisors after training [during quarterly monitoring surveys in Dec 2018] | 0/8 (0%) | 0/6 (0%) | 1/11 (9%) | 1/25 (4%) ^c^ |
|  | Proportion of randomly sampled WDALs reporting that they have received *Andilaye* supportive supervision from HEWs after training [during quarterly monitoring surveys in Dec 2018] | 15/29 (52%) | 5/24 (21%) | 18/43 (42%) | 38/96 (40%) ^c^ |
| **Context** | ^a^ Training of WDALs was planned to primarily be facilitated by HEWs with help from CHC HEWs Supervisors, but the involvement of CHC HEWs Supervisors was minimal in some *kebeles*. Thus, the Ethiopian-based study team assisted trainers and trained WDALs together with HEWs*.*  ^b^ Some *kebeles* added/removed small numbers of WDALs responsible for conducting *Andilaye* counseling visits with caregivers.  ^c^ It was recommended that CHC HEWs Supervisors perform supportive supervision on a routine basis (at least once per month for each HEW), in accordance with the woreda's *Andilaye* action planning document. Similarly, it was recommended that HEWs perform supportive supervision with each WDAL in her catchment area every month, in accordance with the HEW's *Andilaye* action planning document. When conducting supervisory visits, HEWs were trained to use the supportive supervision and on-the-job checklist. However, quarterly monitoring surveys conducted 1-2 months after the review meeting and refresher training of WDALs (Dec 2018) indicated that few HEWs have received supportive supervision and on-the-job training from CHC HEW Supervisors and that few WDALs have received supportive supervision and on-the-job training from HEWs. The review of the importance of providing supportive supervision to both HEWs and WDALs was prioritized during ‘Adaptive management workshop’ (Feb-Mar 2019) and during ‘Skills-based review meetings and refresher trainings for WDALs (round 2)’ (Feb-Mar 2019). | | | | |

(6) Skills-based review meeting and refresher training for WDALs (round 2) (community-level maintenance activity)

| **Process data** | | **Bahir Dar Zuria** | **Fogera** | **Farta** | **Overall** |
| --- | --- | --- | --- | --- | --- |
| **Dose delivered** | Proportion of intervention *kebeles* with activity implemented (Feb-Mar 2019) | 8/8 (100%) | 6/6 (100%) | 11/11 (100%) | 25/25 (100%) |
|  | Proportion of intervention *kebeles* with activity objectives completed as planned (according to activity checklist) | 8/8 (100%) | 6/6 (100%) | 11/11 (100%) | 25/25 (100%) |
|  | Length of activity (one day training) | 2.5 hours | 2.5 hours | 2.66 hours | 2.5 hours |
| **Participation** | Proportion of intervention *kebeles* with HEWs acting as activity training facilitator | 8/8 (100%) | 6/6 (100%) | 11/11 (100%) | 25/25 (100%) |
|  | Proportion of intervention *kebeles* with CHC HEWs Supervisors acting as activity training co-facilitator | 2/8 (25%) | 3/6 (50%) | 4/11 (36%) | 9/25 (36%) ^a^ |
|  | Proportion of WDALs from intervention *kebeles* in attendance during the training (8-10 per *kebele*) | 71/78 (91%) | 53/61 (87%) | 103/107 (96%) | 227/246 (92%) ^b^ |
|  | Proportion of intervention *kebeles* with *Andilaye* trained WDALs | 8/8 (100%) | 6/6 (100%) | 11/11 (100%) | 25/25 (100%) |
| **Dose received** | Proportion of intervention *kebeles* with HEWs reporting that they have received *Andilaye* supportive supervision from CHC HEWs Supervisors after training [during endline surveys in May 2019] | 0/8 (0%) | 3/6 (50%) | 2/11 (18%) | 5/25 (20%) ^c^ |
|  | Proportion of randomly sampled WDALs reporting that they have received *Andilaye* supportive supervision from HEWs after training [during endline surveys in May 2019] | 9/32 (28%) | 5/24 (21%) | 29/44 (66%) | 43/100 (43%) ^c^ |
| **Context** | ^a^ Training of WDALs was planned to primarily be facilitated by HEWs with help from CHC HEWs Supervisors, but the involvement of CHC HEWs Supervisors was minimal in some *kebeles*. Thus, the *Andilaye* team assisted trainers and trained WDALs together with HEWs*.*  ^b^ Some *kebeles* added/removed small numbers of WDALs responsible for conducting *Andilaye* counseling visits with caregivers.  ^c^ It was recommended that CHC HEWs Supervisors perform supportive supervision on a routine basis (at least once per month for each HEW), in accordance with the woreda's *Andilaye* action planning document. Similarly, it was recommended that HEWs perform supportive supervision with each WDAL in her catchment area every month, in accordance with the HEW's *Andilaye* action planning document. When conducting supervisory visits, HEWs were trained to use the supportive supervision and on-the-job checklist. However, endline monitoring surveys conducted 1-2 months after the review meeting and refresher training of WDALs (round 2) indicated that few HEWs have received supportive supervision and on-the-job training from CHC HEW Supervisors and that few WDALs have received supportive supervision and on-the-job training from HEWs. | | | | |

(7) Skills-based refresher training of community conversation facilitators (district-level maintenance activity)

| **Process data** | | **Bahir Dar Zuria** | **Fogera** | **Farta** | **Overall** |
| --- | --- | --- | --- | --- | --- |
| **Dose delivered** | Proportion of study districts with activity implemented (Feb-Apr 2019) | 1/1 (100%) ^a^ | 1/1 (100%) | 1/1 (100%) | 3/3 (100%) |
|  | Proportion of study districts with activity objectives completed as planned (according to activity checklist) | 1/1 (100%) | 1/1 (100%) | 1/1 (100%) | 3/3 (100%) |
|  | Length of activity (one day training) | 8 hours | 8.5 hours | 7.75 hours | 8 hours |
| **Participation** | Total number of participants | 34 | 25 | 46 ^b^ | 105 |
|  | Proportion of study districts with *Woreda* Health Office representative AND Hygiene and Sanitation Officer in attendance | 1/1 (100%) | 0/1 (0%) | 1/1 (100%) | 2/3 (66%) |
|  | Proportion of *Andilaye* community conversation facilitators in attendance | 32/32 (100%) | 24/24 (100%) | 44/44 (100%) | 100/100 (100%) |
|  | Proportion of intervention *kebeles* with at least at least one facilitator per pair of facilitators trained on *Andilaye* community conversations | 8/8 (100%) | 6/6 (100%) | 11/11 (100%) | 25/25 (100%) |
| **Does received** | Proportion of intervention *kebeles* that identified implementation challenges and developed action plan for ways forward for the completion of group-level activities | 8/8 (100%) | 6/6 (100%) | 11/11 (100%) | 25/25 (100%) |
| **Context** | ^a^ Considerable delays were faced when trying to facilitate the Bahir Dar refresher training. While other *woredas* had trainings in February 2019, Bahir Dar Zurias’s refresher training was delayed until early April 2019. These delays resulted from the *woreda’s* prioritization in national health care campaigns in which many trained community conversation facilitators were involved (HEWs, *kebele* administrators, *kebele* managers, etc.). Accordingly, unlike in the original training of community conversation facilitators, all participants were trained together in Bahir Dar Zuria.  ^b^ Two training sessions were conducted in Farta given the high number of intervention *kebeles* and target participants. | | | | |

**Key behavior change activities**

(1) Counseling visits and follow-up barrier planning counseling visits with caregivers (household-level catalyzing and maintenance activity)

| **Process data** | | **Bahir Dar Zuria** | **Fogera** | **Farta** | **Overall** |
| --- | --- | --- | --- | --- | --- |
| **Dose delivered** | Proportion of intervention kebeles with activity implemented (Feb 2018-May 2019) | 8/8 (100%) | 6/6 (100%) | 11/11 (100%) | 25/25 (100%) |
|  | Proportion of respondents from study-enrolled households^¥^ in intervention kebeles reporting they received an Andilaye counseling visit from a WDAL [during endline household visits (May 2019)] | 126/220 (57%) | 75/146 (51%) | 190/299 (64%) | 391/665 (59%) |
|  | Proportion of respondents from study-enrolled households^¥^ in intervention kebeles reporting they received more than one Andilaye counseling visit from a WDAL [during endline household visits (May 2019)] | 83/220 (38%) | 54/146 (37%) | 149/299 (50%) | 286/665 (43%) |
|  | Average number of Andilaye counseling visits reported [of non-WDAL households^¥^ reporting at least one Andilaye counseling visit during endline household visits (May 2019)] | 298/126 (2.4) | 189/75 (2.5) | 559/190 (2.9) | 1,046/391 (2.7) ^a^ |
| **Participation** | Proportion of study-enrolled households in intervention kebeles with an Andilaye goal card hung in the house [observed during endline household visits (May 2019)] | 142/241 (59%) | 95/171 (56%) | 246/331 (74%) | 483/743 (65%) ^b^ |
|  | Proportion of respondents from study-enrolled households^¥^ in intervention kebeles that could identify the WDAL responsible for conducting their Andilaye counseling visits [during endline household visits (May 2019)] | 140/220 (64%) | 88/146 (60%) | 211/299 (71%) | 439/665 (66%) |
| **Dose received** | Proportion of respondents from study-enrolled households in intervention kebeles reporting they set household goals or incremental improvements [of non-WDAL households reporting at least one Andilaye counseling visit during endline household visits (May 2019)] | 86/126 (68%) | 57/75 (76%) | 138/190 (73%) | 281/391 (72%) ^b^ |
|  | Proportion of respondents from study-enrolled households in intervention kebeles reporting they discussed barriers to the goals or incremental improvements [of non-WDAL households reporting at least one Andilaye counseling visit during endline household visits (May 2019)] | 75/126 (60%) | 52/75 (70%) | 131/190 (69%) | 258/391 (66%) ^b^ |
|  | Proportion of respondents from study-enrolled households in intervention kebeles reporting they discussed solutions to the barriers of the goals or incremental improvements [of non-WDAL households reporting at least one Andilaye counseling visit during endline household visits (May 2019)] | 72/126 (57%) | 46/75 (61%) | 124/190 (65%) | 242/391 (62%) ^b^ |
| **Context** | ^¥^ Excluding study-enrolled households that were residents of caregivers who were trained as WDALs responsible for conducting the Andilaye counselling visits with cargivers (Bahir Dar Zuria, n=5; Fogera, n=15; Farta, n=22)  ^a^ Per protocol, WDALs were to act as the primary counselor and visit each household in her catchment area to conduct an Andilaye household counselling visit about once per month, with each visit lasting around 30 minutes. WDALs from all intervention kebeles implemented Andilaye counseling visits with caregivers. However, no kebele had WDALs conducting monthly counseling visits according to caregivers from study-enrolled households in intervention kebeles surveyed during endline. Of households reporting at least one visit (n=391), the average number of visits was 2-3 during the 14-15 months of implementation (i.e., since the initial trainings in January-February 2018). This suggests visits were likely only conducted following each round of WDAL training (n=3) for a majority of WDALs.  ^b^ All behavior change techniques designed into the Andilaye counseling visits with caregivers (e.g., inter-personal counseling on action planning, barrier identification and planning; goal setting, commitment, and self-regulation), were critical to the Andilaye intervention. However, of the households reporting at least one counseling visit (n=391), only 72% reported that they set household goals or incremental improvements, and two-thirds reported that they identified barriers, and their WDAL provided counseling on how to plan for, cope with, and overcome barriers – suggesting that some households receiving visits were not exposed to the intended intervention, as designed. | | | | |

(2) Community mobilization and commitment event (community-level catalyzing activity)

| **Process data** | | **Bahir Dar Zuria** | **Fogera** | **Farta** | **Overall** |
| --- | --- | --- | --- | --- | --- |
| **Dose delivered** | Proportion of intervention kebeles with activity implemented (Mar-April 2018) | 8/8 (100%) | 6/6 (100%) | 11/11 (100%) | 25/25 (100%) |
|  | Proportion of intervention kebeles with activity objectives completed as planned (according to activity checklist) | 8/8 (100%) | 6/6 (100%) | 11/11 (100%) | 25/25 (100%) ^a^ |
|  | Average length of activity (partial day event) | 2.33 hours | 2 hours | 2 hours | 2 hours |
| **Participation** | Estimated average number of adult community members in attendance per event | 355 | 298 | 279 | 309 ^b^ |
|  | Proportion of respondents from study-enrolled households in intervention kebeles reporting being aware of the Andilaye community mobilization and commitment event [during quarterly monitoring visits (July and Dec 2018)] | 35/226 (15%) | 47/172 (27%) | 77/305 (25%) | 159/703 (22%) ^b^ |
|  | Proportion of respondents from study-enrolled households in intervention kebeles reporting attending the Andilaye community mobilization and commitment event [during quarterly monitoring visits (July and Dec 2018)] | 29/226 (13%) | 43/172 (25%) | 56/305 (18%) | 128/703 (18%) ^b^ |
|  | Proportion of intervention kebeles with ALL coordinating committee members in attendance | 3/8 (38%) | 1/6 (17%) | 5/11 (45%) | 9/25 (36%) ^c^ |
|  | Proportion of coordinating committee members in attendance | 16/34 (47%) | 15/27 (56%) | 29/45 (64%) | 60/106 (57%) ^c^ |
|  | Proportion of intervention kebeles with ALL masters of ceremony in attendance | 1/8 (13%) | 3/6 (50%) | 3/11 (27%) | 9/25 (36%) ^c^ |
|  | Proportion of masters of ceremony in attendance | 14/22 (64%) | 11/15 (73%) | 20/31 (65%) | 45/68 (66%) ^c^ |
|  | Proportion of intervention kebeles with ALL HEWs in attendance | 5/8 (63%) | 3/6 (50%) | 7/11 (64%) | 15/25 (60%) |
|  | Proportion of intervention kebeles with at least one HEW in attendance | 7/8 (88%) | 6/6 (100%) | 11/11 (100%) | 24/25 (96%) ^d^ |
|  | Proportion of intervention kebeles with ALL Andilaye trained WDALs in attendance | 1/8 (13%) | 3/6 (50%) | 5/11 (45%) | 9/25 (36%) |
|  | Proportion of Andilaye trained WDALs in attendance | 39/73 (53%) | 41/56 (73%) | 84/110 (76%) | 164/229 (72%) |
| **Dose received** | Proportion of intervention kebeles that determined practices no longer deemed to be acceptable by the community at the end of the event | 8/8 (100%) | 6/6 (100%) | 10/11 (91%) | 24/25 (96%) |
|  | Proportion of intervention kebeles that determined improved behaviors at the end of the event | 8/8 (100%) | 6/6 (100%) | 9/11 (82%) | 23/25 (92%) |
|  | Proportion of intervention kebeles that determined regulations for monitoring the by-laws at the end of the event | 6/8 (75%) | 6/6 (100%) | 9/11 (82%) | 21/25 (84%) ^e^ |
| **Context** | ^a^ The objectives were completed as planned, however, slight difference in the quality of event performance groups hired for skits and music during the events were noticed. Four performance groups were hired to perform in the 25 intervention kebeles. The two performance groups hired from Farta who performed in kebeles in Farta and Fogera performed better than the two performance groups who were hired from Bahir Dar Zuria. Since equal payment was decided for all performance groups hired, it was not possible to get best performance groups in Bahir Dar Zuria because of more opportunities for performance groups in Bahir Dar Zuria than in Farta (i.e., higher quality Bahir Dar Zuria performance groups were more expensive).  ^b^ Mobilizing community members to come to the event presented a challenge in participation. Mobilization and overall community planning of the event was tasked to the coordinating committee and masters of ceremony identified during the ‘Whole System in the Room’ activity. The payment (per diem) issue associated with the ‘Whole System in the Room’ might have impacted the motivation of some coordinating committee members and masters of ceremony to follow through with activities identified in the action plan.  ^c^ Many absent coordinating committee members and masters of ceremony were reportedly engaged in other government meetings during the day of the event. The Ethiopian-based study team was in attendance for all events and efforts by the team were made by to assist coordinating committee members and masters of ceremony that were in attendance of the event to mobilize and conduct the event.  ^d^ HEW turnover presented a challenge in HEW attendance in one intervention kebele.  ^e^ Setting community by-laws and determining regulations for monitoring the by-laws was a challenge in some kebeles. In few kebeles, HEWs expressed their concern on determining regulations considering the current political situation (i.e., recent declarations of states of emergency) and public protests in the region in the last few years. In one kebele, it was noted that practices no longer deemed acceptable by the community and practices that need to be improved were already determined in previous WASH meetings and the community leaders stated there is no need to do the same thing again; the community leaders agreed to write the already set by-laws on the banner later the day. In another kebele, the kebele administrator and manager were not willing to lead a discussion for developing by-laws and left the event before the pledge and commitment event. Two other kebele, agreed to determine regulations for monitoring the by-laws in the following weeks of the event. | | | | |

| **Process data** | | **Bahir Dar Zuria** | **Fogera** | **Farta** | **Overall** |
| --- | --- | --- | --- | --- | --- |
| **Dose delivered** | Proportion of intervention *kebeles* with activity implemented (Oct 2018-May 2019) | 6/8 (75%) | 6/6 (100%) | 11/11 (100%) | 23/25 (92%) ^a^ |
|  | Proportion of intervention *kebeles* with activity implemented as designed (i.e., at least two groups of community members attending community conversation sessions for all three behavioral themes) (Oct 2018-May 2019) [reported by facilitators at endline (May 2019)] | 2/8 (25%) | 5/6 (83%) | 7/11 (64%) | 14/25 (56%) |
| **Participation** | Proportion of respondents from study-enrolled households in intervention *kebeles* reporting that they have heard about the *Andilaye* community conversations [during quarterly monitoring household visits (May 2019)] | 63/225 (28%) | 76/161 (47%) | 190/321 (57%) | 329/707 (46%) |
|  | Proportion of respondents from study-enrolled households in intervention *kebeles* reporting attending at least one *Andilaye* community conversations [during endline household visits (May 2019)] | 30/225 (13%) | 48/161 (30%) | 122/321 (38%) | 200/707 (28%) ^b^ |
| **Context** | ^a^ Community conversation facilitators from two *kebeles* in Bahir Dar Zuria reported that they had not conducted community conversations within the 8 months since the initial training. Facilitators reported competing priorities that require the immediate action of facilitators. Overall, facilitators reported that punctuality and motivation of participants as a challenge in conducting community conversations. During refresher trainings (Feb-Apr 2019), many facilitators suggested addressing these issues by encouraging participation within the community by-laws as well as working with *kebele* officials to mobilize households.  ^b^ While the reported attendance of respondents is low, it should be noted that community conversations were designed to address a variety of participants beyond primary caregivers (such as, husbands/fathers, community or religious leaders/elders, youths/students, *kebele* officials/administrators, health development leaders, etc.). | | | | |

(3) Community conversations and follow up community conversations (group-level catalyzing and maintenance activity)

**Supplemental Table 4.** *Andilaye* Trial indicators used to assess WASH behavior

| **Indicators** | **Overall** | | **Intervention** | | **Control** | |  |  |
| --- | --- | --- | --- | --- | --- | --- | --- | --- |
| **Sanitation** |  | |  | |  | |  |  |
| **Household latrine coverage** | **N** | **%** | **N** | **%** | **N** | **%** | **PR (95% CI)^a^** | **PD (95% CI)^b^** |
| Households with at least one latrine | 1472 | 61.6 | 743 | 61.2 | 729 | 62.0 | 0.99 (0.82, 1.21) | -0.0036 (-0.12, 0.12) |
| Households with improved latrine | 1467 | 32.5 | 741 | 34.6 | 726 | 30.3 | 1.13 (0.81, 1.59) | 0.0408 (-0.07, 0.15) |
| Households with latrine with smooth and cleanable slab/floor | 1471 | 14.8 | 743 | 16.3 | 728 | 13.3 | 1.19 (0.70, 2.03) | 0.0263 (-0.05, 0.11) |
| Households with fully constructed latrine | 1471 | 30.9 | 742 | 33.0 | 729 | 28.7 | 1.15 (0.86, 1.54) | 0.0440 (-0.46, 0.13) |
| Percent of household latrines that were fully constructed | 906 | 50.1 | 454 | 53.4 | 452 | 46.2 | **1.18 (1.01, 1.38)** | **0.0820 (0.01, 0.16)** |
| **Household latrine characteristics** |  |  |  |  |  |  |  |  |
| Stagnant water present over the floor / latrine slab | 906 | 17.0 | 455 | 16.3 | 451 | 17.7 | 0.92 (0.66, 1.28) | -0.0137 (-0.07, 0.04) |
| Pan, slab, or floor was discolored (e.g., yellow, green) | 904 | 59.7 | 454 | 58.4 | 450 | 61.1 | 0.97 (0.88, 1.08) | -0.0175 (-0.08, 0.04) |
| Presence of flies in latrine | 906 | 83.2 | 455 | 81.8 | 451 | 84.7 | 0.97 (0.89, 1.06) | -0.0226 (-0.95,0.05) |
| Presence of drop hole cover in the latrine | 906 | 14.1 | 455 | 18.2 | 451 | 10.0 | **1.77 (1.19, 2.63)** | **0.0789 (0.02, 0.14)** |
| Among those with a drop hole, a cover was situated over drop hole | 130 | 67.7 | 85 | 70.6 | 45 | 62.2 | 1.09 (0.88, 1.36) | 0.0579 (-0.08, 0.20) |
| Presence of cleaning agents for washing latrine | 906 | 5.7 | 455 | 8.4 | 451 | 3.1 | 2.46 (0.76, 7.96) | 0.0419 (-0.02, 0.11) |
| Presence of feces on floor / slab or other place in the latrine | 906 | 50.6 | 455 | 50.1 | 451 | 51.0 | 1.02 (0.90, 1.15) | 0.0080 (-0.05, 0.07) |
| Evidence latrine is used for storage or other non-sanitation-related purpose | 906 | 7.6 | 455 | 7.7 | 451 | 7.5 | 0.93 (0.49, 1.75) | -0.0054 (-0.05, 0.04) |
| Presence of well-worn path to latrine | 906 | 95.1 | 455 | 95.4 | 451 | 94.9 | 1.01 (0.97, 1.05) | -0.0047 (-0.03, 0.04) |
| Presence of fresh feces on / in the pit or pan | 906 | 73.3 | 455 | 73.6 | 451 | 73.0 | 1.01 (0.90, 1.12) | 0.0034 (-0.07, 0.08) |
| Pit is full or close to being full | 905 | 12.2 | 454 | 11.7 | 451 | 12.6 | 0.92 (0.57, 1.49) | -0.0098 (-0.68, 0.05) |
| Presence of anal cleansing item inside or near latrine | 906 | 40.3 | 455 | 39.1 | 451 | 41.5 | 0.93 (0.77, 1.12) | -0.0287 (-1.02, 0.04) |
| Presence of odor from stool or urine in the latrine | 906 | 84.0 | 455 | 81.8 | 451 | 86.3 | 0.96 (0.87, 1.04) | -0.0381 (-0.11, 0.04) |
| Presence of leaves, spider webs, rubbish, other dirt in latrine | 906 | 20.3 | 455 | 18.5 | 451 | 22.2 | 0.85 (0.60, 1.12) | -0.0348 (-0.11, 0.04) |
| Wet latrine floor | 906 | 43.8 | 455 | 40.7 | 451 | 47.0 | 0.85 (0.68, 1.07) | -0.0675 (-0.16, 0.03) |
| Presence of water available near or inside latrine for hand washing | 906 | 12.3 | 455 | 18.0 | 451 | 6.4 | **2.28 (1.08, 4.81)** | 0.0885 (-0.01, 0.18) |
| Presence of cleansing agent near or inside latrine for hand washing | 905 | 2.8 | 454 | 5.3 | 451 | 0.2 | **21.67 (2.17, 215.98)** | 0.0698 (-0.04, 0.18) |
| Water available inside or near latrine for flushing or self-cleansing | 906 | 4.5 | 455 | 6.6 | 451 | 2.4 | **2.35 (1.05, 5.28)** | 0.0356 (-0.01, 0.08) |
| **Household latrine facility operation and maintenance** |  |  |  |  |  |  |  |  |
| Latrine cleaned during the last seven days | 904 | 23.2 | 454 | 25.6 | 450 | 20.9 | 1.02 (0.65, 1.60) | 0.0039 (-0.95, 1.03) |
| Household has added or improved anything on this latrine since its original construction | 899 | 16.5 | 453 | 17.2 | 446 | 15.7 | 1.08 (0.71, 1.65) | 0.0120 (-0.06, 0.08) |
| Have ever fixed anything that became broken, damaged, our worn out on this latrine since its original construction | 900 | 23.1 | 453 | 23.4 | 447 | 22.8 | 1.02 (0.76, 1.37) | 0.0049 (-0.06, 0.07) |
| Is your latrine facility working (operating) correctly now? | 907 | 93.4 | 455 | 94.1 | 452 | 92.7 | 1.02 (0.98, 1.05) | 0.0140 (-0.02, 0.05) |
| Facility observed to require obvious repair | 906 | 75.3 | 455 | 70.1 | 451 | 80.5 | **0.88 (0.78, 0.99)** | **-0.0985 (-0.19, -0.01)** |
| The latrine was observed to be serviceable | 906 | 78.8 | 455 | 80.0 | 451 | 77.6 | 1.05 (0.93, 1.18) | 0.0391 (-0.05, 0.13) |
| Cleaning agents for washing latrine were observed inside or near the latrine | 906 | 5.7 | 455 | 8.4 | 451 | 3.1 | 2.46 (0.76, 7.96) | 0.0419 (-0.03, 0.11) |
| Presence of feces on floor, slab, or other place in the latrine aside from the pit | 906 | 50.1 | 455 | 50.1 | 451 | 51.0 | 1.02 (0.90, 1.15) | 0.0080 (-0.05, 0.07) |
| **Latrine utilization** | **N** | **mean (SE)** | **N** | **mean (SE)** | **N** | **mean (SE)** | **-** | **difference (95% CI)^e^** |
| Given you have a latrine, number of households that used this household latrine in last 7 days | 905 | 1.63 (0.09) | 454 | 1.54 (0.09) | 451 | 1.72 (0.14) | - | -0.1144 (-0.42, 0.19) |
| Given household has a latrine, number of people who used this latrine from another household during last 7 days, not including your household members | 902 | 0.91 (0.14) | 454 | 0.70 (0.16) | 448 | 1.11 (0.22) | - | -0.3990 (-0.85, 0.05) |
|  | **N** | **%** | **N** | **%** | **N** | **%** | **PR (95% CI)^a^** | **PD (95% CI)^b^** |
| Given you have a latrine, household members have used this latrine for 3 or more days during the last 7 days | 905 | 90.8 | 455 | 92.1 | 450 | 89.6 | 1.04 (0.99, 1.10) | 0.0378 (-0.13, 0.09) |
| Respondent defecated in any latrine during last 2 days | 1469 | 45.5 | 740 | 45.8 | 729 | 45.3 | 1.01 (0.79, 1.29) | 0.0025 (-0.11, 0.12) |
| Respondent always exclusively used a latrine for defecation during last 7 days | 1472 | 46.4 | 743 | 53.2 | 729 | 54.1 | 0.99 (0.79, 1.24) | -0.0044 (-0.12, 0.12) |
| Respondent’s primary place of defecation changes over the course of the year | 1470 | 27.3 | 742 | 28.8 | 728 | 25.7 | 1.16 (0.90, 1.50) | 0.0406 (-0.03, 0.11) |
| Head of household defecated in any latrine during last 2 days | 900 | 57.7 | 486 | 57.6 | 414 | 57.7 | 1.01 (0.83, 1.23) | 0.0053 (-0.11, 0.12) |
| Head of household always exclusively used a latrine for defecation during last 7 days | 1002 | 34.8 | 529 | 36.5 | 473 | 33.0 | 1.07 (0.79, 1.47) | 0.0270 (-0.09, 0.15) |
| Head of household’s primary place of defecation changes over the course of the year | 1205 | 25.9 | 624 | 25.5 | 581 | 26.3 | 0.96 (0.71, 1.30) | -0.0101 (-0.09, 0.07) |
| Ages 4-17 defecated in any latrine during last 2 days | 2991 | 52.1 | 1538 | 52.7 | 1453 | 51.2 | 1.01 (0.83, 1.20) | 0.0046 (-0.10, 0.11) |
| Ages 4-17 always exclusively used a latrine for defecation during last 7 days | 2842 | 38.9 | 1447 | 42.6 | 1385 | 35.0 | 1.15 (0.89, 1.50) | 0.0573 (-0.05, 0.16) |
| Ages 4-17 primary place of defecation changes over the course of the year | 3532 | 23.5 | 1778 | 24.5 | 1754 | 22.4 | 1.08 (0.83, 1.41) | 0.0170 (-0.05, 0.08) |
| Safely disposed of child feces | 777 | 38.9 | 401 | 36.7 | 376 | 41.2 | 0.96 (0.69, 1.32) | -0.0171 (-0.15, 0.11) |
| **Open defecation practices** |  |  |  |  |  |  |  |  |
| Respondent’s primary place of defecation was OD during last 2 days | 1472 | 39.5 | 743 | 40.2 | 729 | 38.8 | 1.05 (0.76, 1.45) | 0.0196 (-0.11, 0.15) |
| Respondent openly defecated during last 2 days | 1472 | 44.9 | 743 | 45.2 | 729 | 44.9 | 1.03 (0.78, 1.36) | 0.0121 (-0.11, 0.14) |
| Respondent openly defecated in or near surface water | 660 | 10.9 | 335 | 10.2 | 325 | 11.7 | 0.93 (0.50, 1.72) | -0.0083 (-0.08, 0.06) |
| Respondent urinated in/or near surface water | 1466 | 7.2 | 739 | 6.5 | 727 | 8.0 | 0.82 (0.46, 1.48) | -0.0140 (-0.06, 0.03) |
| Head of household’s primary place of defecation was OD during last 2 days | 1146 | 40.3 | 600 | 39.7 | 546 | 41.0 | 0.95 (0.68, 1.31) | -0.0226 (-0.15, 0.11) |
| Head of household openly defecated during last 2 days | 992 | 64.7 | 527 | 62.6 | 465 | 67.1 | 0.98 (0.80, 1.21) | -0.0126 (-0.14, 0.12) |
| Head of household openly defecated in or near surface water during last 2 days | 371 | 12.4 | 199 | 11.6 | 172 | 13.3 | 1.07 (0.48, 2.40) | 0.0084 (-0.09, 0.10) |
| Head of household urinated in/near surface water | 704 | 7.7 | 387 | 6.7 | 317 | 8.8 | 0.82 (0.35, 1.94) | -0.0145 (-0.08, 0.05) |
| Ages 4-17 primary place of defecation was OD during last 2 days | 3494 | 44.3 | 1757 | 42.9 | 1737 | 45.7 | 0.97 (0.72, 1.29) | -0.0149 (-0.14, 0.11) |
| Ages 4-17 openly defecated during last 2 days | 2921 | 58.8 | 1489 | 55.2 | 1432 | 62.6 | 0.95 (0.77, 1.16) | -0.0310 (-0.15, 0.09) |
| Ages 4-17 openly defecated in or near surface water during last 2 days | 1326 | 18.4 | 629 | 14.5 | 697 | 22.0 | 0.73 (0.39, 1.37) | -0.0540 (-0.17, 0.06) |
| Ages 4-17 urinated in/near surface water | 2480 | 11.7 | 1273 | 9.0 | 1207 | 14.5 | 0.66 (0.35, 1.24) | -0.05 (-0.12, 0.03) |
| Evidence of open defecation (i.e., human feces) in/near household compound | 1472 | 45.2 | 743 | 43.5 | 729 | 47.1 | 0.95 (0.78, 1.15) | -0.0258 (-0.11, 0.06) |
| **Personal hygiene** |  |  |  |  |  |  |  |  |
| **Washing station coverage** |  |  |  |  |  |  |  |  |
| Reported HH hand or facewashing station(s) | 1472 | 98.0 | 743 | 98.3 | 729 | 97.7 | 1.01 (0.99, 1.02) | 0.0076 (-0.01, 0.02) |
| Observed HH hand or facewashing station(s) with water | 1472 | 19.9 | 743 | 19.5 | 729 | 20.3 | 0.96 (0.72, 1.26) | -0.0090 (-0.06, 0.05) |
| Observed HH hand or facewashing station(s) with soap (%) | 1472 | 2.2 | 743 | 2.6 | 729 | 1.9 | 1.34 (0.44, 4.13) | 0.0066 (-0.02, 0.04) |
| **Handwashing practices** |  |  |  |  |  |  |  |  |
| Yesterday, the index child's hands were washed | 1428 | 98.5 | 724 | 99.0 | 704 | 97.9 | **1.01 (1.00, 1.02)** | **0.0116 (0.00, 0.02)** |
| The last time the index child's hands were washed, soap/ash/or soapy water used | 1398 | 44.8 | 714 | 46.5 | 684 | 43.0 | 1.08 (0.89, 1.30) | 0.0329 (-0.05, 0.12) |
| The last time the index child defecated, he/she cleaned hands with water and soap, soapy water, or ash | 1410 | 41.8 | 713 | 43.9 | 697 | 39.6 | 1.12 (0.92, 1.35) | 0.0461 (-0.04, 0.13) |
| Yesterday, the respondent washed his/her hands with water | 1469 | 98.0 | 742 | 98.3 | 727 | 97.8 | 1.01 (0.99, 1.02) | 0.0057 (-0.01, 0.02) |
| The last time the respondent washed he/she used soap/ash/soapy water | 1468 | 44.0 | 740 | 46.0 | 728 | 42.0 | 1.09 (0.91, 1.31) | 0.0390 (-0.04, 0.12) |
| The last time the respondent defecated, he/she cleaned hands with water and soap | 1463 | 49.0 | 738 | 51.9 | 725 | 46.1 | 1.13 (0.94, 1.35) | 0.0585 (-0.03, 0.15) |
| The last time the respondent prepared food, he/she cleaned hands with water and soap before beginning food preparations | 1403 | 51.0 | 700 | 53.6 | 703 | 48.5 | 1.11 (0.95, 1.29) | 0.0513 (-0.03, 0.13) |
| **Hand cleanliness** |  |  |  |  |  |  |  |  |
| Respondent’s finger nails clean on left hand | 1472 | 73.6 | 743 | 74.6 | 729 | 72.7 | 1.03 (0.96, 1.11) | 0.0227 (-0.03, 0.08) |
| Respondent’s finger nail clean on right hand | 1472 | 73.4 | 743 | 73.6 | 729 | 73.1 | 1.01 (0.94, 1.09) | 0.0101 (-0.04, 0.65) |
| Respondent’s finger pads clean on left hand | 1472 | 62.1 | 743 | 63.3 | 729 | 60.9 | 1.05 (0.95, 1.15) | 0.0275 (-0.03, 0.09) |
| Respondent’s Finger pads clean on right hand | 1472 | 62.2 | 743 | 62.7 | 729 | 61.7 | 1.03 (0.93, 1.13) | 0.0157 (-0.04, 0.08) |
| Respondent’s palm clean on left hand | 1472 | 57.5 | 743 | 59.0 | 729 | 56.1 | 1.05 (0.94, 1.18) | 0.0292 (-0.04, 0.09) |
| Respondent’s palm clean on right hand | 1472 | 57.9 | 743 | 59.1 | 729 | 56.6 | 1.05 (0.94, 1.17) | 0.0265 (-0.04, 0.09) |
| Index child’s finger nails clean on left hand | 1024 | 84.1 | 502 | 83.3 | 522 | 84.9 | 0.96 (0.91, 1.02) | -0.0342 (-0.08, 0.14) |
| Index child’s finger nail clean on right hand | 1024 | 83.9 | 502 | 83.1 | 522 | 84.7 | 0.96 (0.90, 1.02) | -0.0324 (-0.08, 0.02) |
| Index child’s finger pads clean on left hand | 1024 | 80.6 | 502 | 79.9 | 522 | 81.2 | 0.98 (0.91, 1.05) | -0.0169 (-0.07, 0.03) |
| Index child’s finger pads clean on right hand | 1024 | 80.4 | 502 | 79.7 | 522 | 81.0 | 0.98 (0.91, 1.05) | -0.0179 (-0.07, 0.03) |
| Index child’s palm clean on left hand | 1024 | 78.9 | 502 | 78.9 | 522 | 78.9 | 0.99 (0.93, 1.05) | -0.0101 (-0.06, 0.04) |
| Index child’s palm clean on right hand | 1024 | 78.6 | 502 | 78.5 | 522 | 78.7 | 0.98 (0.92, 1.05) | -0.0120 (-0.06, 0.04) |
| **Facewashing practices** |  |  |  |  |  |  |  |  |
| Yesterday, the index child's face was cleaned | 1433 | 98.1 | 723 | 97.9 | 710 | 98.3 | 0.99 (0.98, 1.01) | -0.0054 (-0.02, 0.01) |
| Yesterday, after index child’s face was washed it was it wiped dry with a cloth | 1325 | 37.1 | 665 | 38.1 | 660 | 36.2 | 1.04 (0.90, 1.21) | 0.0160 (-0.04, 0.07) |
| Yesterday, the respondent cleaned his/her face | 1470 | 98.0 | 742 | 97.6 | 728 | 98.4 | 0.99 (0.97, 1.01) | -0.006 (-0.03, 0.01) |
| Yesterday, the respondent wiped his/her face dry with a cloth such as a towel or apron | 1393 | 39.1 | 702 | 40.9 | 691 | 37.3 | 1.09 (0.09, 1.27) | 0.0326 (-0.03, 0.10) |
| **Face cleanliness** |  |  |  |  |  |  |  |  |
| Ocular discharge is present (all children ages 1-9 years) | 1696 | 28.7 | 822 | 26.9 | 874 | 30.4 | 0.88 (0.68, 1.15) | -0.0369 (-0.11, 0.04) |
| Wet nasal discharge is present (all children ages 1-9 years) | 1696 | 38.2 | 822 | 37.0 | 874 | 39.4 | 0.94 (0.78, 1.13) | -0.0244 (-0.09, 0.05) |
| Dry nasal discharge is present (all children ages 1-9 years) | 1696 | 44.0 | 822 | 42.7 | 874 | 45.2 | 0.97 (0.81, 1.16) | -0.0157 (-0.10, 0.06) |
| Dirt/dust/other debris is present (all children ages 1-9 years) | 1696 | 50.0 | 822 | 50.5 | 874 | 49.5 | 1.03 (0.89, 1.20) | 0.0157 (-0.06, 0.09) |
| Ocular discharge is present (index child) | 1024 | 29.8 | 502 | 27.5 | 522 | 32.0 | 0.88 (0.68, 1.15) | -0.0369 (-0.11, 0.04) |
| Wet nasal discharge is present (index child) | 1024 | 39.8 | 502 | 37.7 | 522 | 42.0 | 0.94 (0.78, 1.13) | -0.0244 (-0.09, 0.05) |
| Dry nasal discharge is present (index child) | 1024 | 45.6 | 502 | 43.2 | 522 | 47.9 | 0.97 (0.81, 1.16) | -0.0157 (-0.05, 0.06) |
| Dirt/dust/other debris is present (index child) | 1024 | 53.0 | 502 | 52.2 | 522 | 52.9 | 1.03 (0.89, 1.20) | 0.0157 (-0.06, 0.09) |
|  | **N** | **mean (SE)** | **N** | **mean (SE)** | **N** | **mean (SE)** |  | **difference (95% CI)^e^** |
| Number of times a fly land on the index child's face during a 1 minute observation | 1024 | 3.3 (0.21) | 502 | 3.2 (0.33) | 522 | 3.4 (0.26) | - | -0.15 (-0.90, 0.60) |
| **Household environmental sanitation** |  |  |  |  |  |  |  |  |
| **Animal husbandry and hygiene practices** | **N** | **%** | **N** | **%** | **N** | **%** | **PR (95% CI)^a^** | **PD (95% CI)^b^** |
| Respondent has animal herding or other animal husbandry responsibilities | 1472 | 87.4 | 743 | 88.3 | 729 | 86.4 | 1.02 (0.96, 1.09) | 0.0196 (-0.04, 0.07) |
| Head of household has animal herding or other animal husbandry responsibilities | 1243 | 91.4 | 639 | 92.5 | 604 | 90.2 | 1.05 (0.99, 1.11) | 0.0421 (-0.01, 0.09) |
| Observed animal feces present in the compound | 1472 | 82.3 | 743 | 82.2 | 729 | 82.4 | 1.01 (0.92, 1.11) | 0.0071 (-0.07, 0.08) |
| Animal feces/waste not left out in open in compound | 1472 | 53.8 | 743 | 56.4 | 729 | 51.2 | 1.10 (0.95, 1.28) | 0.0516 (-0.03, 0.13) |
| **Solid waste management** |  |  |  |  |  |  |  |  |
| Solid waste was not observed to have been left out in the open | 1472 | 31.1 | 743 | 34.6 | 729 | 27.6 | 1.26 (0.93, 1.69) | 0.0705 (-0.02, 0.17) |
